# Validity of the Cold Pressor Test and Pain Sensitivity Questionnaire via online self-administration

**DOI:** 10.1101/19011775

**Authors:** Matthew H. McIntyre, 23andMe Research Team, Achim Kless, Peter Hein, Mark Field, Joyce Y. Tung

## Abstract

To determine the feasibility of complex home-based phenotyping, 1,876 research participants from the customer base of 23andMe participated in an online version of a Pain Sensitivity Questionnaire (PSQ) as well as a cold pressor test (CPT) which is used in clinical assessments of pain. Overall our online version of the PSQ performed similarly to the original pen-and-paper version. Construct validity of the PSQ total was demonstrated by internal consistency and consistent discrimination between more and less painful items. Criterion validity was demonstrated by correlation with pain sensitivity as measured by the cold pressor test. Within the same cohort we performed a cold pressor test using a layperson description and household equipment. Comparison with published reports from controlled studies revealed similar distributions of cold pain tolerance times (i.e., time elapsed before removing the hand from the water). Of those who elected to participate in the CPT, a large majority of participants did not report issues with the test procedure or noncompliance to the instructions (97%). We confirmed a large sex difference in CPT thresholds in line with published data, such that women removed their hands from the water at a median of 54.2 seconds, with men lasting for a median time of 82.7 seconds (Kruskal-Wallis statistic, p < 0.0001), but other factors like age or current pain treatment were at most weakly associated, and inconsistently between men and women. We introduce a new paradigm for performing pain testing, called testing@home, that, in the case of cold nociception, showed comparable results to studies conducted under controlled conditions and supervision of a health care professional.

**Summary:** Research paradigms employing home-based phenotyping are feasible, with both questionnaires and self-administration of a well-established experimental human pain model yielding similar results compared to controlled settings.

## Introduction

Drug development in pain indications has a higher-than-average attrition rate. One possible way to increase success is to identify promising targets based on genetic links between target and disease ^13^. However, discovering new genetic links to pain requires large cohorts ^26^, which may be difficult to obtain in using phenotypes obtained under controlled settings (e.g. clinical trials). In order to enable collection of larger samples, we investigated pain phenotyping in a large cohort of subjects using an internet and home-based approach.

Deep pain phenotyping of subjects is a key prerequisite to identify subgroups for a pain indication. In a laboratory quantitative sensory testing (QST) protocol, subjects may be exposed to multiple types of pain stimulus (heat, cold, pressure, etc.). Here we present the results from 1,876 participants asked to perform the cold pressor test (CPT) on themselves at home, via a layperson description, using household equipment, and guided by a web-based workflow. In addition to the CPT, participants self-administered an online version of the Pain Sensitivity Questionnaire (PSQ) and reported about their history of treatment for pain.

It is well established that individuals experience pain differently ^2,5^. A similar sensory input caused by experimental or clinical pain can lead to vastly differing ratings of the pain experience by different subjects, and it is most likely that this is caused by individual differences in both peripheral and central processing of that sensory input. This has significant implications for patients and clinicians who may struggle to establish a treatment that is effective in reducing pain experience (as indicated by pain ratings), but also for drug development where the differences described above introduce another layer of variability, making it more difficult to identify true drug effects. Although the interindividual differences in pain processing are well known, there is only limited data available describing this variability in larger populations.

The CPT was developed to measure autonomous responses in the cardiovascular system ^7^. The test usually consists of immersion of one hand in ice water for a specified amount of time ^12^, which induces both pain and a response of the autonomous nervous system. The CPT has been widely adopted as a model for nociceptive pain, and for opioids it is established as a surrogate of clinical efficacy ^19^. As such, this test has been used mostly in small populations from clinical trials with two notable exceptions: cohorts from Haifa, Israel, ^14,15,23^ and unpublished data, and Tromsø, Norway ^8^, both summarized by Treister et al. ^22^. The studies conducted in Haifa included 648 people. The Tromsø Study included 10,486 people enriched for those 40-42 or 60-87 years old. That study found much lower pain sensitivity than seen in the participants from Haifa, in that most participants left their hands in the cold water for longer than 100 seconds. However, they found that large group of participants with chronic pain removed their hands from the water sooner, indicating lower pain tolerance.

The PSQ consists of 14 imagined painful situations and 3 non-painful control situations, and subjects are asked to rate their painfulness on a 0-10 numeric rating scale, originally in German ^16^. Ruscheweyh et al. showed that the PSQ demonstrated strong internal consistency, demonstrated by high Cronbach’s α of both the PSQ-total score and two derived sub-factors labeled as sensitivity to “minor” and “moderate” pain. Moreover, they reported evidence of criterion validity demonstrated by correlation with subjective pain experienced from a range of stimuli, including pinprick, pressure, phasic and tonic heat and cold, and the cold pressor test. However, PSQ-total score did not correlate with response thresholds to any stimulus, including time to withdraw one’s hand from cold water in the CPT. A validation in chronic pain patients ^16^, and a separate English language validation of the PSQ ^18^ have been published. However, the number of subjects from which PSQ data are published is limited (406 subjects in ^16^, 319 subjects in ^17^, 136 subjects in ^18^).

Pain sensitivity measured via the PSQ may be associated with a history of chronic pain. In a second study, Ruscheweyh et al. reported significantly elevated PSQ scores in 134 chronic pain patients as compared to 185 healthy controls. A subgroup of 46 chronic pain patients were given experimental pain testing, not including the CPT but including pain ratings, but not thresholds, from another tonic cold stimulus. In that subgroup, there was a strong correlation between PSQ and tonic cold stimulus pain rating ^17^.

Applying pain ratings to studies requiring large samples, such as genome-wide association studies ^26^. While large studies of pain sensitivity have been conducted in relatively controlled settings ^6,8,10^, self-administration allow for large studies to be conduct more quickly and at less expense. Here, we investigate whether online, self-administered versions of the PSQ and CPT demonstrate similar construct and criterion validity as observed in both pen-and-paper and laboratory protocols.

## Methods

### Tools/measures - Web-based phenotyping

23andMe is a direct to consumer personal genomics company with a research platform that allows participants to complete questionnaires for research purposes. We added online versions CPT consent and instructions, and questionnaires, to the research area of the 23andMe Personal Genetics Service website (www.23andme.com). Ultimately, 1,876 participants who were at least 20 years old, and consented to participate in the research, completed the questionnaires followed by the CPT. In addition to the PSQ questionnaire, we asked about history of diagnosis with, and treatment for, acute and chronic pain-related conditions. As our pain history questionnaire was unexpectedly long for many participants, the 1,876 participants who completed the activities reflect substantial attrition from the approximately 10,000 who started the sequence.

### Subjects

We recruited participants for both the questionnaires and CPT using email and by participation of research participants on the 23andMe website in the US. Participants provided informed consent online according to a protocol approved by Ethical & Independent Review Services, a private institutional review board (OHRP/FDA registration number IRB00007807, study number 10044-11), which included separate consent for the CPT. Participants with pain conditions were eligible for this study, though they were excluded from the original validation study of the PSQ by Ruscheweyh et al. ^16^. Exclusions for participation are described in Table 1. Participants with self-reported cardiovascular disorders (e.g. high blood pressure, heat diseases, dysrhythmia), a history of Raynaud’s phenomenon, any neurological disorders, and/or pregnancy were not recruited and advised not to participate (Table 1) to minimize the risk of adverse events during the CPT procedure. Especially because no researcher was present in this at-home testing who could provide assistance in such cases we used this as an additional safety precaution. This resulted in the exclusion of participants with some chronic pain conditions, like migraine, but not others, like back pain. Ultimately, of the 1,876 participants, 181 (11.2%) reported being treated in the past 4 months with prescription medication for an acute pain condition and another 78 (4.2%) for a chronic pain condition. Those treated for both chronic and acute pain conditions were classified as having acute pain.

**Table 1.** Study exclusion criteria, as represented to participants.

|  |
| --- |
| High blood pressure |
| Heart disease |
| Dysrhythmia |
| Any other cardiovascular disorder |
| History of Raynaud's phenomenon |
| History of fainting or seizures |
| History of frostbite |
| An open cut, sore, or bone fracture on or near your non-dominant hand (the one you do not usually write with) |
| Any neurological disorder |
| Are pregnant or think you might be pregnant |

### Differences to supervised CPT

In contrast to a controlled clinical or laboratory setting, in our study the CPT was done by the participants themselves at home using a lay description of the procedure (Figure 1). Two primary outcomes were assessed: cold pain threshold and cold pain tolerance. Cold pain threshold was the time to the first report of pain and cold tolerance the tome time to removal of the hand from the water. We additionally asked about maximum pain intensity, but our measure of pain intensity differs from some other laboratory protocols. First, we presented an 11-point visual analog scale immediately after the completion of the task, rather than periodically during the task itself. This was required to minimize the number of simultaneous tasks for participants to perform. Second, we only asked about the maximum pain felt at the end of the test. While at least one large study has measure pain intensity in a similar way ^14,15^, others score periodic measurements by averaging over all pain intensity measurements and imputing the maximum score 10, for any intervals following the removal of the hand ^8^. Studies may also use both methods ^1^.

**Figure 1.**
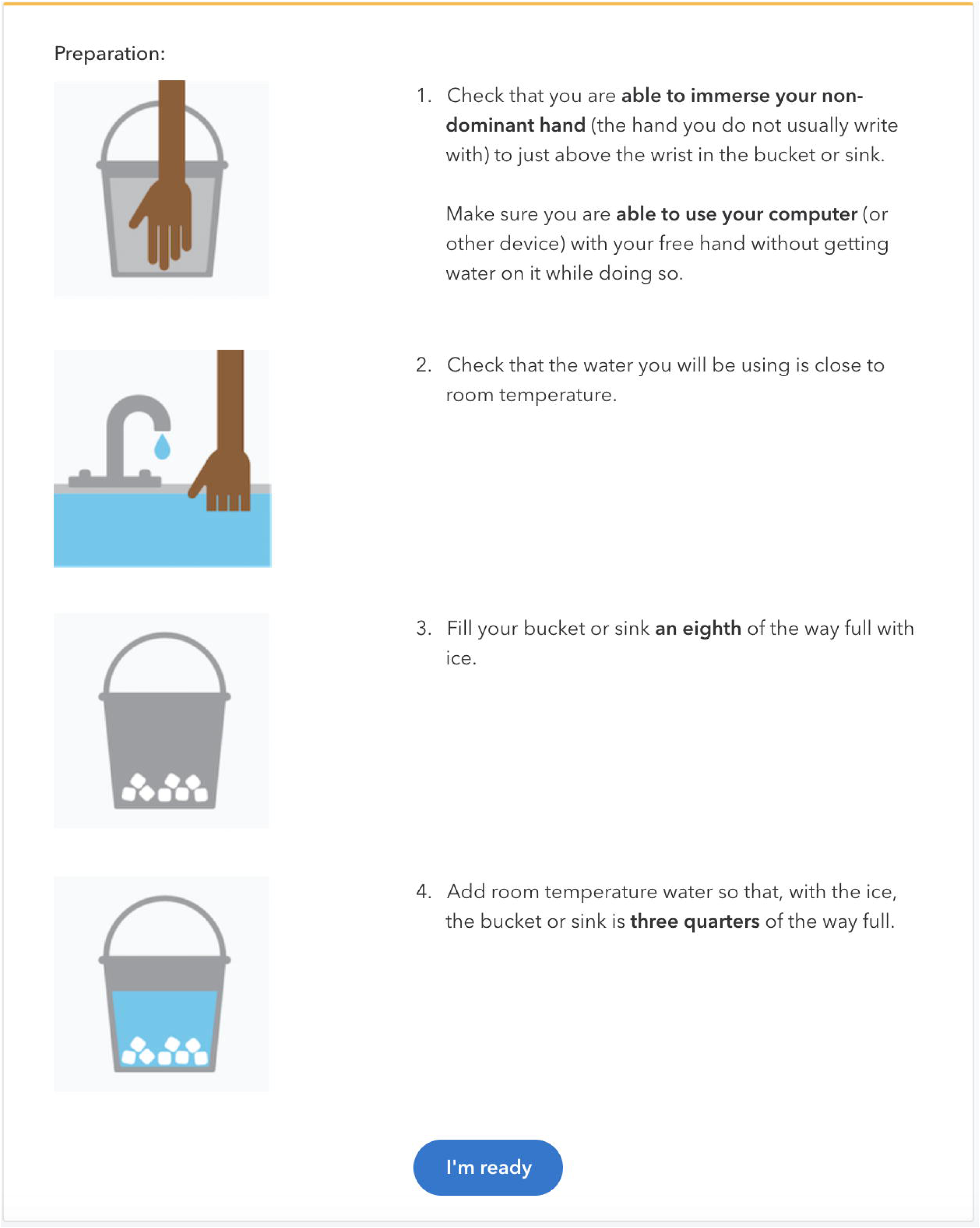
Web-based workflow of cold pressor test.

We analyzed the PSQ by calculating the PSQ total score and by factor analysis. In addition to comparing derived factors with published reports, we assessed the extent to which each participant’s rating correlated with the factor loadings, reflecting the extent to which they rated experiences more indicative of pain sensitivity more highly than ones less indicative, regardless of the average level of painfulness report. Finally, we compared PSQ scores with CPT thresholds and pain intensity ratings.

### Questionnaires

In a sequence of two questionnaires, the first included the English-language version of the PSQ and additional questions about the participant’s own memory of painful experiences. The second questionnaire included questions about history of pain conditions and related medications.

Causes of chronic pain included self-reported low back pain; complex regional pain syndrome; joint pain; diabetic neuropathy; endometriosis; cancer; other internal pain not caused by endometriosis or cancer; shingles, cold sores, or herpes; trigeminal neuralgia; migraine; non-migraine headaches that occurred more than half of the days in any given month; pain following amputation; or other pain after injury or surgery that lasted more than 3 months. Acute pain included dental pain, pain after injury or surgery that lasted less than 3 months, or other pain that lasted less than 3 months. Participants were classified as having current acute pain if they had been treated with a prescription pain medication for an acute condition in past 4 months. Participants without current acute pain were classified as having current chronic pain if they had been treated with a prescription pain medication for a chronic condition in the past 4 months. Otherwise, they were classified with no current pain condition.

### Cold pressor test

Participants with a history of migraine and a number of other chronic conditions that might be directly exacerbated by the activity were not invited to participate in the CPT in order to ensure their safety, while participants with other chronic pain conditions, including chronic back pain, were invited. Moreover, participants reporting neurological or temperature-triggered conditions (e.g. migraine, history of syncope, or Raynaud’s phenomenon) or current injuries to their non-dominant hands at the time of recruitment were ineligible to review the supplementary CPT consent document and to participate. Participants were asked to prepare their own bath of ice water at home, and to keep their non-dominant hand submerged to the wrist for no more than 150 seconds. The amount of ice in the instructions (1/8 of the container size) would not fully melt at ambient water and air temperatures, and would remain floating at the top of the water. However, if participants did not add enough ice or added warm water and the ice did melt, temperatures would have risen from about 0° at the beginning to a higher temperature at the end of the test. This differs from many laboratory protocols in which refrigeration and water circulation are used to keep the temperature at 2-5° for the duration of the test.

### Data analysis

To test the psychometric validity of the online English-language PSQ ^18^, we compared psychometric properties with those of the original validation of the German-language version of the PSQ ^16^. We computed the same measures as described in the original study of the German language version: PSQ-total (all items considered painful), PSQ-minor (the least painful items: 14, 3, 6, 12, 11, 10, and 7, ordered from least to most painful), PSQ-moderate (8, 15, 2, 16, 17, 1, 4 ordered from least to most painful), and calculated Cronbach’s α for each measure, a measure of internal consistency.

To further assess construct validity, we conducted factor analysis of the PSQ items. Factor analysis of the PSQ items by Ruscheweyh et al. ^16^ yielded two factors corresponding to minor and moderate causes of pain. On the basis of this result, they proposed a PSQ-total score (the mean of all painful items) and PSQ-minor and PSQ-moderate subscales (the means of the respective items). Our comparisons with Ruscheweyh et al. ^16^ include those between (1) item-level means scores, (2) mean scores for the PSQ-total and subscales, (3) varimax factor loading, (4) item correlations with PSQ-total, (5) correlation of CPT pain threshold, tolerance, and intensity with PSQ-total to assess criterion validity of both the PSQ and CPT. We also compare the distribution of CPT pain thresholds with those found in previous studies, including Ruscheweyh et al. ^16^. Following the recommendation of Treister et al. ^22^, we used the Kruskal-Wallis rank test to examine differences between, sex, age, and pain history group in CPT thresholds and pain intensity.

In addition to these validation steps, we also investigated whether participants varied in the accuracy. We defined accuracy as the within-person correlation of PSQ pain ratings with the loadings on the first principal component. Those who rated the items that were generally considered more painful as more painful, or vice-versa, would have high accuracy scores.

## Results

### Pain Sensitivity Questionnaire

Internal consistencies (Cronbach’s α) were similar to Ruscheweyh et al. ^16^ for PSQ total (ours 0.93, theirs 0.92), PSQ minor (ours 0.84, theirs 0.81), and PSQ moderate (ours 0.90, theirs 0.91). Table 2 gives mean scores for the whole group as well as scores stratified by sex, age, and pain history. Items for the PSQ-moderate (1, 2, 4, 8, 15, 16, and 17) were each rated as more painful than items for the PSQ-minor (3, 6, 7, 10, 11, 12, and 14) in both the present study and Ruscheweyh et al. ^16^ (see Table 3 and 4). Three items (5, 9, 13) that describe normally non-painful situations have been excluded.

**Table 2.** Mean PSQ total, minor and moderate scores for the total study population as well as stratified scores for sex, age, and current pain.

|  | Count | PSQ total<br>mean (SD) | PSQ minor<br>mean (SD) | PSQ moderate<br>mean (SD) |
| --- | --- | --- | --- | --- |
| Ruscheweyh et al. (2009) | 354 | 3.6 (1.2) | 2.5 (1.1) | 4.7 (1.6) |
| 23andMe/Grünenthal online cohort<br>Total | 1876 | 3.1 (1.4) | 2.2 (1.2) | 4.0 (1.6) |
| SEX: Male | 679 | 3.0 (1.3) | 2.1 (1.1) | 3.9 (1.6) |
| SEX: Female | 1197 | 3.2 (1.4) | 2.3 (1.3) | 4.0 (1.7) |
| AGE: 20-29 | 356 | 3.3 (1.3) | 2.4 (1.2) | 4.2 (1.5) |
| AGE: 30-39 | 426 | 3.1 (1.3) | 2.2 (1.2) | 4.0 (1.6) |
| AGE: 40-49 | 297 | 3.0 (1.4) | 2.1 (1.3) | 3.9 (1.6) |
| AGE: 50-59 | 285 | 3.1 (1.4) | 2.2 (1.3) | 4.1 (1.7) |
| AGE: 60-69 | 332 | 2.9 (1.3) | 2.0 (1.2) | 3.8 (1.7) |
| AGE: >70 | 180 | 3.1 (1.4) | 2.1 (1.2) | 4.1 (1.7) |
| PAIN: Acute | 181 | 3.2 (1.5) | 2.2 (1.3) | 4.1 (1.8) |
| PAIN: Chronic | 78 | 3.2 (1.5) | 2.3 (1.4) | 4.0 (1.7) |
| PAIN: None | 1617 | 3.1 (1.3) | 2.2 (1.2) | 4.0 (1.6) |

**Table 3.** Ratings for individual PSQ items and factor structures observed in online sample vs. original sample. Items used to calculate the PSQ moderate subscale are indicated in **bold**. Others comprise the moderate subscale.

| Item | Mean<br>(SD) | Factor loading<br>Online PSQ-E<br>n=1,876 |  | Factor loading<br>Ruscheweyh et al. [12]<br>Table 2<br>n = 354 |  | Correlation<br>with PSQ<br>total<br>Online | Correlation<br>with PSQ<br>total<br>Ruscheweyh<br>et al. [12] |
| --- | --- | --- | --- | --- | --- | --- | --- |
|  |  | Moderate<br>(2) | Minor<br>(1) | Moderate<br>(1) | Minor<br>(2) |  |  |
| **1 bump shin** | **4.3 (2.0)** | **0.83** | **0.19** | **0.77** | **0.12** | **0.66** | **0.62** |
| **2 burn tongue** | **3.4 (1.9)** | **0.80** | **0.24** | **0.78** | **0.21** | **0.68** | **0.69** |
| 3 sore muscles | 2.3 (1.7) | 0.62 | 0.33 | 0.12 | 0.78 | 0.61 | 0.51 |
| **4 trap finger** | **4.5 (2.1)** | **0.77** | **0.34** | **0.77** | **0.28** | **0.75** | **0.73** |
| 6 sunburn shoulders | 1.8 (1.5) | 0.33 | 0.55 | 0.08 | 0.77 | 0.57 | 0.47 |
| 7 graze knee | 3.1 (1.8) | 0.49 | 0.58 | 0.42 | 0.53 | 0.70 | 0.59 |
| **8 bite cheek** | **3.4 (2.0)** | **0.58** | **0.54** | **0.72** | **0.36** | **0.77** | **0.74** |
| 10 cut finger | 2.5 (1.8) | 0.25 | 0.76 | 0.41 | 0.65 | 0.70 | 0.67 |
| 11 prick finger | 2.0 (1.5) | 0.30 | 0.76 | 0.48 | 0.56 | 0.74 | 0.66 |
| 12 hands in snow | 2.6 (2.1) | 0.19 | 0.71 | 0.32 | 0.51 | 0.65 | 0.51 |
| 14 strong hand<br>shake | 1.1 (1.5) | 0.17 | 0.53 | 0.25 | 0.53 | 0.53 | 0.47 |
| **15 hot handle** | **4.6 (2.1)** | **0.43** | **0.63** | **0.71** | **0.35** | **0.72** | **0.73** |
| **16 crush foot** | **4.2 (2.1)** | **0.53** | **0.58** | **0.77** | **0.30** | **0.77** | **0.75** |
| 17 funny bone | 3.5 (2.1) | 0.56 | 0.52 | 0.79 | 0.31 | 0.74 | 0.76 |

**Table 4.** Imagined situations for pain ratings used in the questionnaire (Ruscheweyh et al. ^16^).

|  |  |
| --- | --- |
| 1 | Imagine you bump your shin badly on a hard edge, for example, on the edge of a glass coffee table. |
| 2 | Imagine you burn your tongue on a very hot drink. |
| 3 | Imagine your muscles are slightly sore as the result of physical activity. |
| 4 | Imagine you trap your finger in a drawer. |
| 5 | Imagine you take a shower with lukewarm water. |
| 6 | Imagine you have mild sunburn on your shoulders. |
| 7 | Imagine you grazed your knee falling off your bicycle. |
| 8 | Imagine you accidentally bite your tongue or cheek badly while eating. |
| 9 | Imagine walking across a cool tiled floor with bare feet. |
| 10 | Imagine you have a minor cut on your finger and inadvertently get lemon juice in the wound. |
| 11 | Imagine you prick your fingertip on the thorn of a rose. |
| 12 | Imagine you stick your bare hands in the snow for a couple of minutes or bring your hands in contact with snow for some time, for example, while making snowballs. |
| 13 | Imagine you shake hands with someone who has a normal grip. |
| 14 | Imagine you shake hands with someone who has a very strong grip. |
| 15 | Imagine you pick up a hot pot by inadvertently grabbing its equally hot handles. |
| 16 | Imagine you are wearing sandals and someone with heavy boots steps on your foot. |
| 17 | Imagine you bump your elbow on the edge of a table (“funny bone”). |

While we found only one principal component with an eigenvalue over 1, the second was close to 1 (7.25 and 0.97), so we compared with the two-factor solution presented by Ruscheweyh et al. ^16^. These two components explain about 59% of the total variance, compared to 55% in Ruscheweyh et al. ^16^. Varimax rotation results in two factors with a similar structure to that observed by Ruscheweyh et al. ^16^, as shown in Table 3, but with several notable differences. Our Factor 1 loads mostly minor causes of pain, similar to Factor 2 in Ruscheweyh et al. ^16^ and our Factor 2 loads more moderate causes of pain. The biggest difference is seen for item 3 (“Imagine your muscles are slightly sore as the result of physical activity”), which loaded on the minor pain factor in Ruscheweyh et al. ^16^, but more heavily load on the moderate pain factor in our results, despite being rated as less painful. Conversely, items 15 (“Imagine you pick up a hot pot by inadvertently grabbing its equally hot handles”) and 16 (“Imagine you wear sandals and someone with heavy boots steps on your foot”) more heavily loaded the moderate pain factor in Ruscheweyh et al. ^16^, but more heavily load the minor pain factor in our results, despite being rated as relatively painful.

Items correlated similarly with the PSQ total score in the two studies (correlation of correlations = 0.92, see Table 3). Pain attributed to a strong handshake (#14) was the weakest correlate of total PSQ score in both studies, and, as a non-painful item, is also not a component of the PSQ scores. The three strongest correlates with PSQ total score in both studies were pain attributed to hitting one’s “funny bone” (#17), to having one’s foot stepped on (#16), and to biting one’s cheek (#8).

### Cold pressor test

Cold pain tolerance distribution is presented in Figure 2. This figure also shows results from two other large cohorts (data from ^22^), obtained in a controlled laboratory setting with a standard methodology (e.g. use of a circulating, refrigerated water bath kept at a constant temperature).

**Figure 2.**
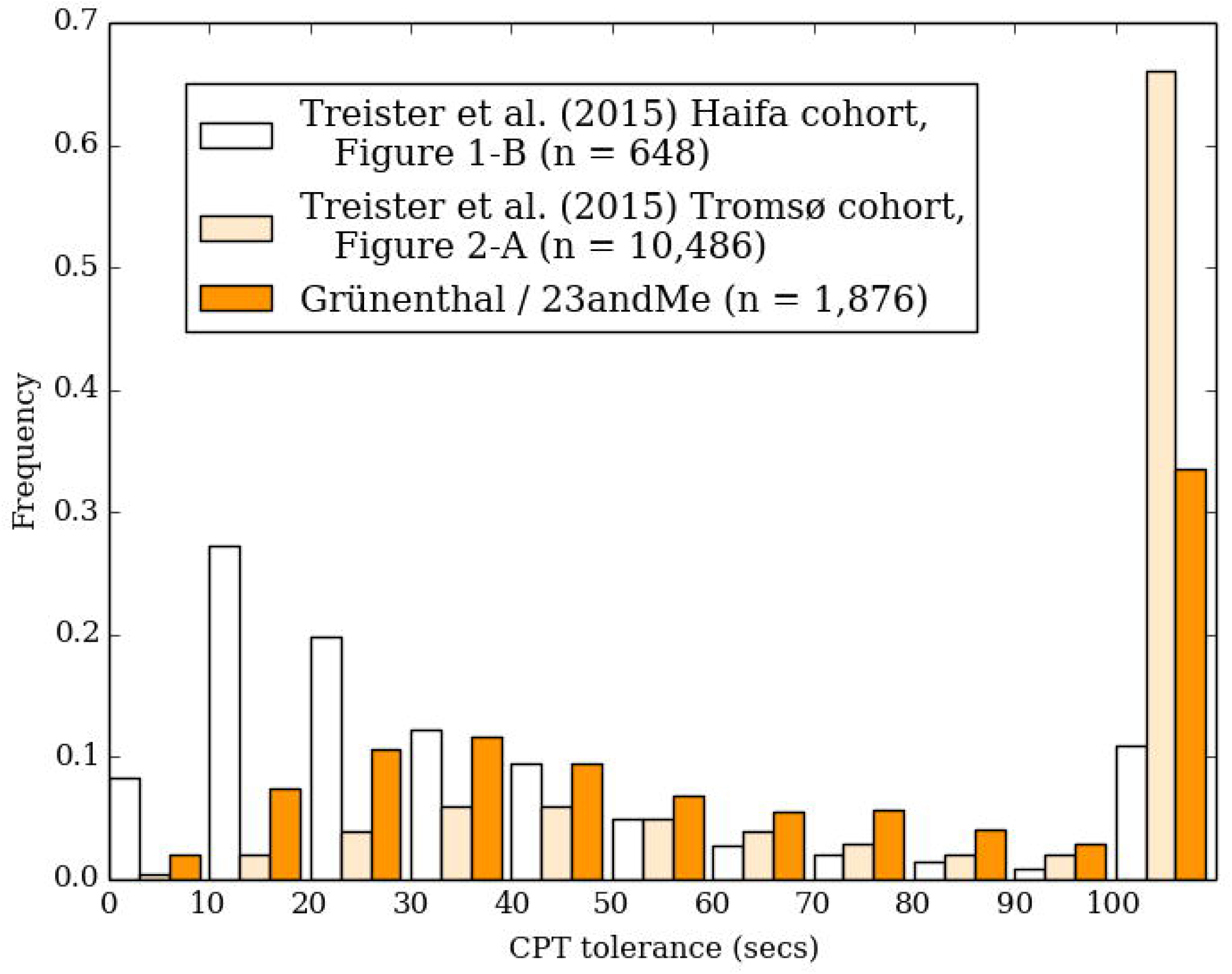
Comparison of the pain tolerance time (time to withdrawal of hand from cold water) of the CPT in our study compared to the data from two other large cohorts (as reviewed by Treister et al. ^22^).

CPT tolerance time distributions differed substantially by sex (Kruskal-Wallis statistic = 71, p<0.0001, Figure 4), but not by age (Kruskal-Wallis statistic = 13, p=0.0241, Figure 5) or pain history (Kruskal-Wallis statistics = 4, p=0.1591, Figure 6). Women report a median tolerance of 54.2 seconds (IQR 30.4 – 116.5), which was 31.0 seconds earlier than the median of 82.7 seconds (IQR 43.6 – 150.0) reported by men.

**Figure 3.**
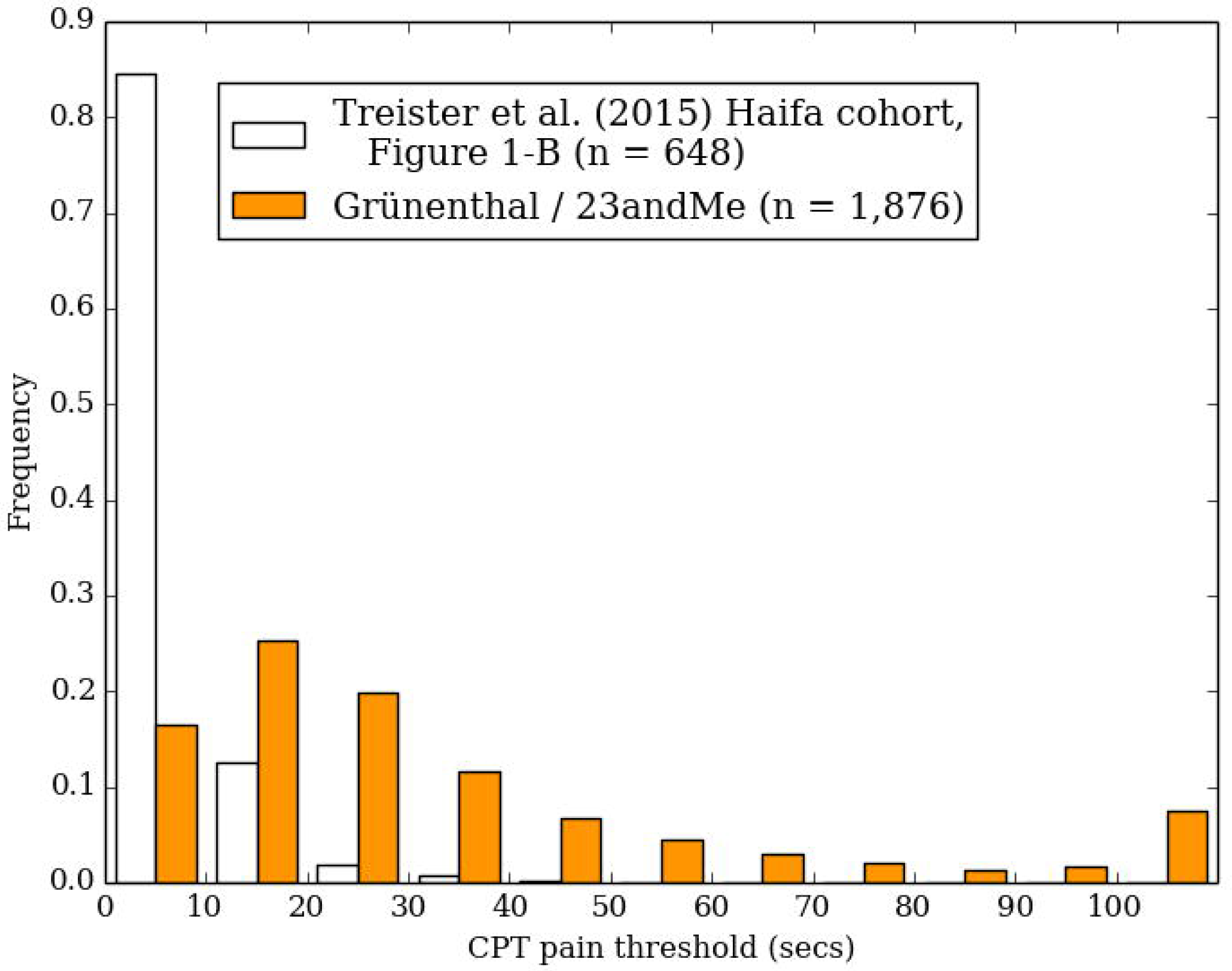
Comparison of the pain threshold time (time to initial report of pain) of the CPT in our study compared to the data from two other large cohorts (as reviewed by Treister et al. ^22^).

**Figure 4.**
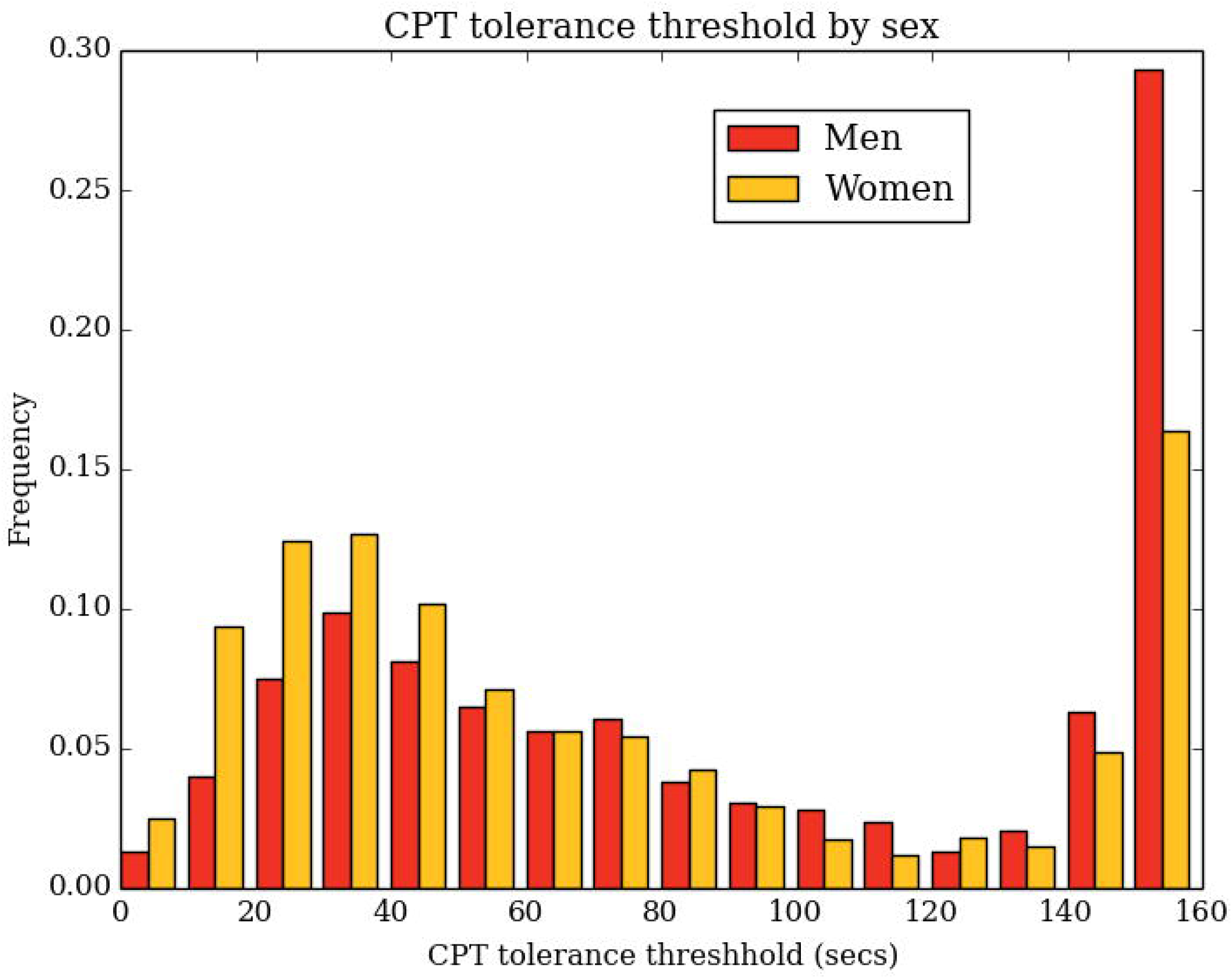
Distributions of CPT tolerance times by sex.

**Figure 5.**
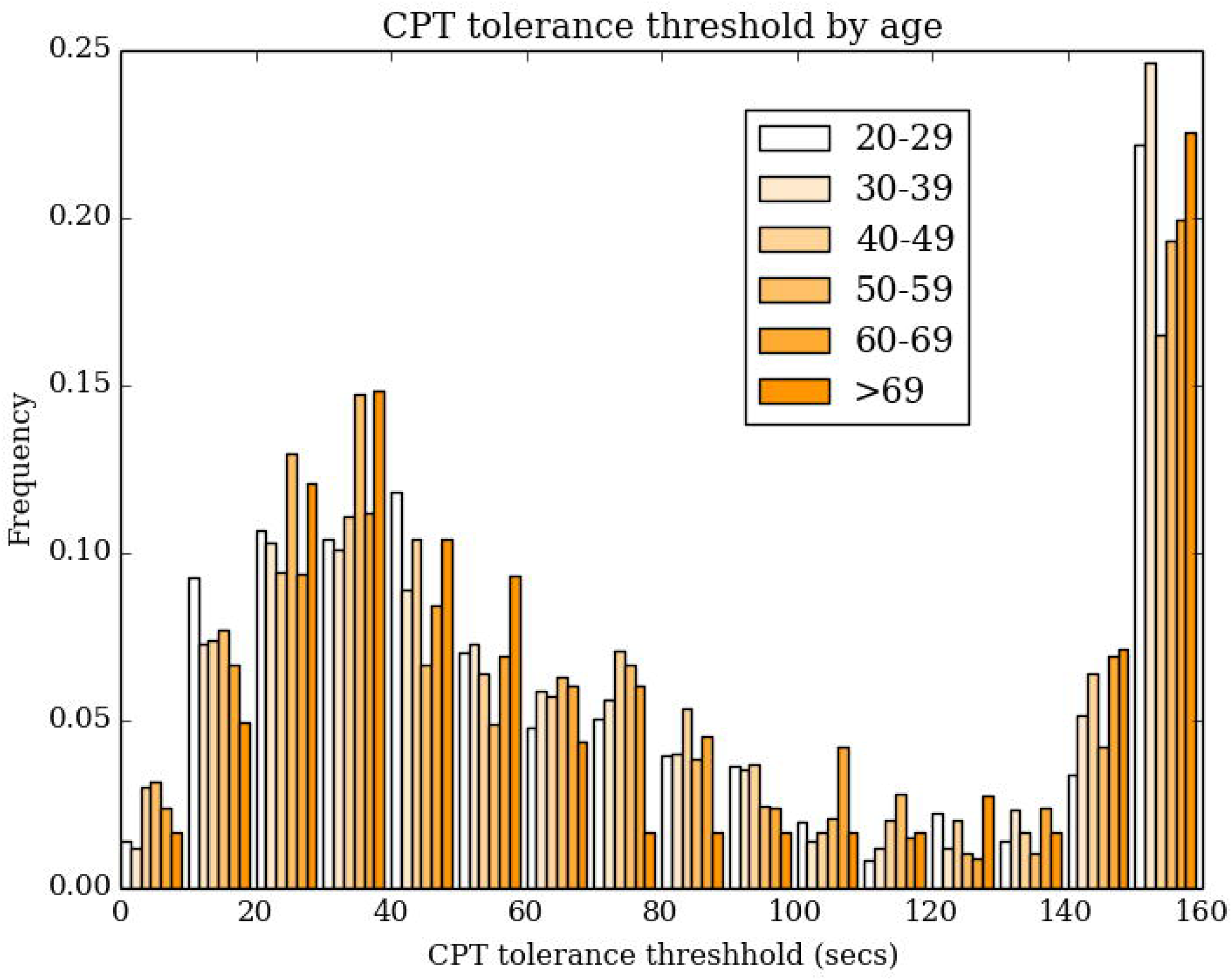
Distributions of CPT tolerance times by age.

**Figure 6.**
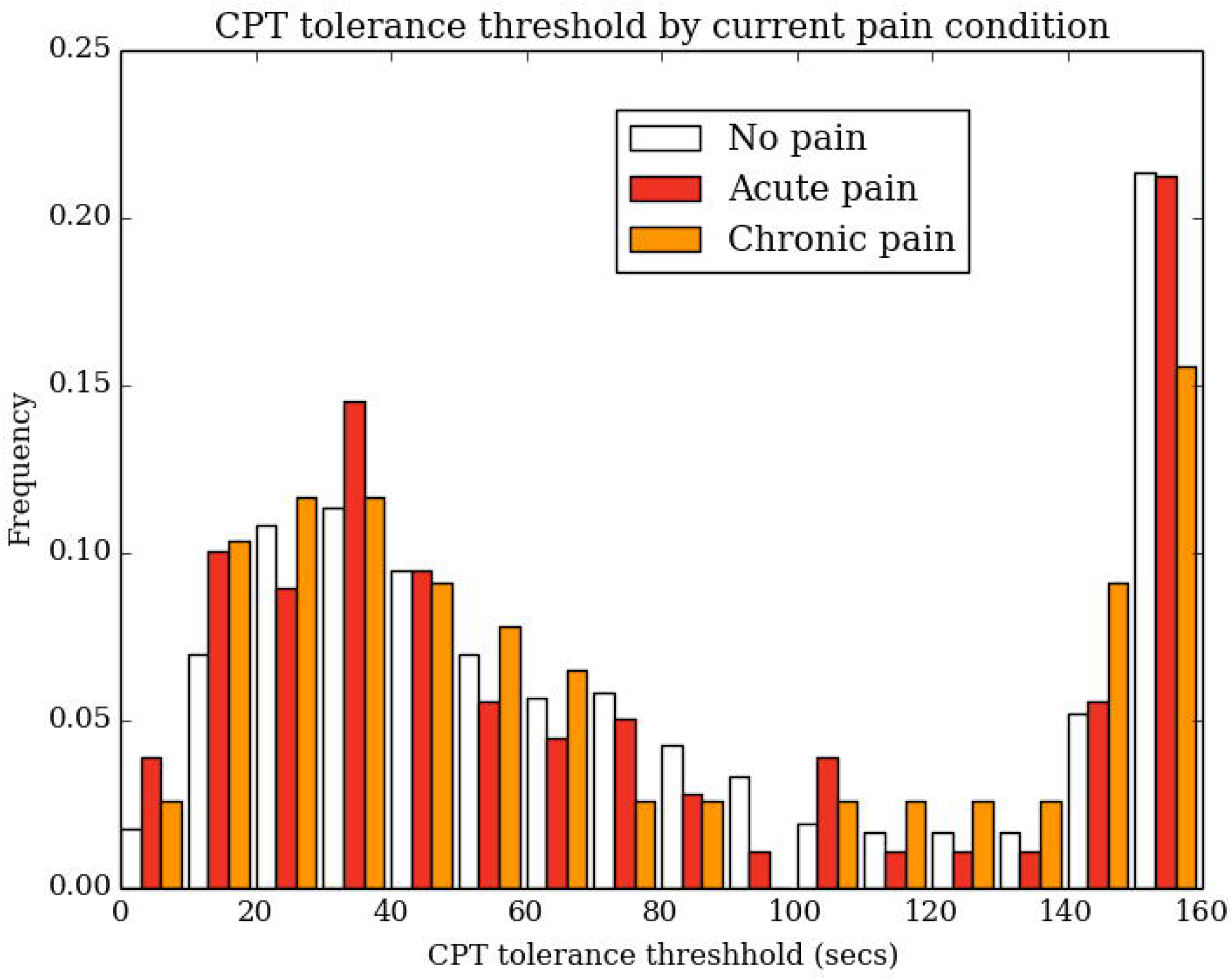
Distributions of CPT tolerance times by current pain status among 1,993 participants who completed by the CPT and a pain history questionnaire.

**Figure 7.**
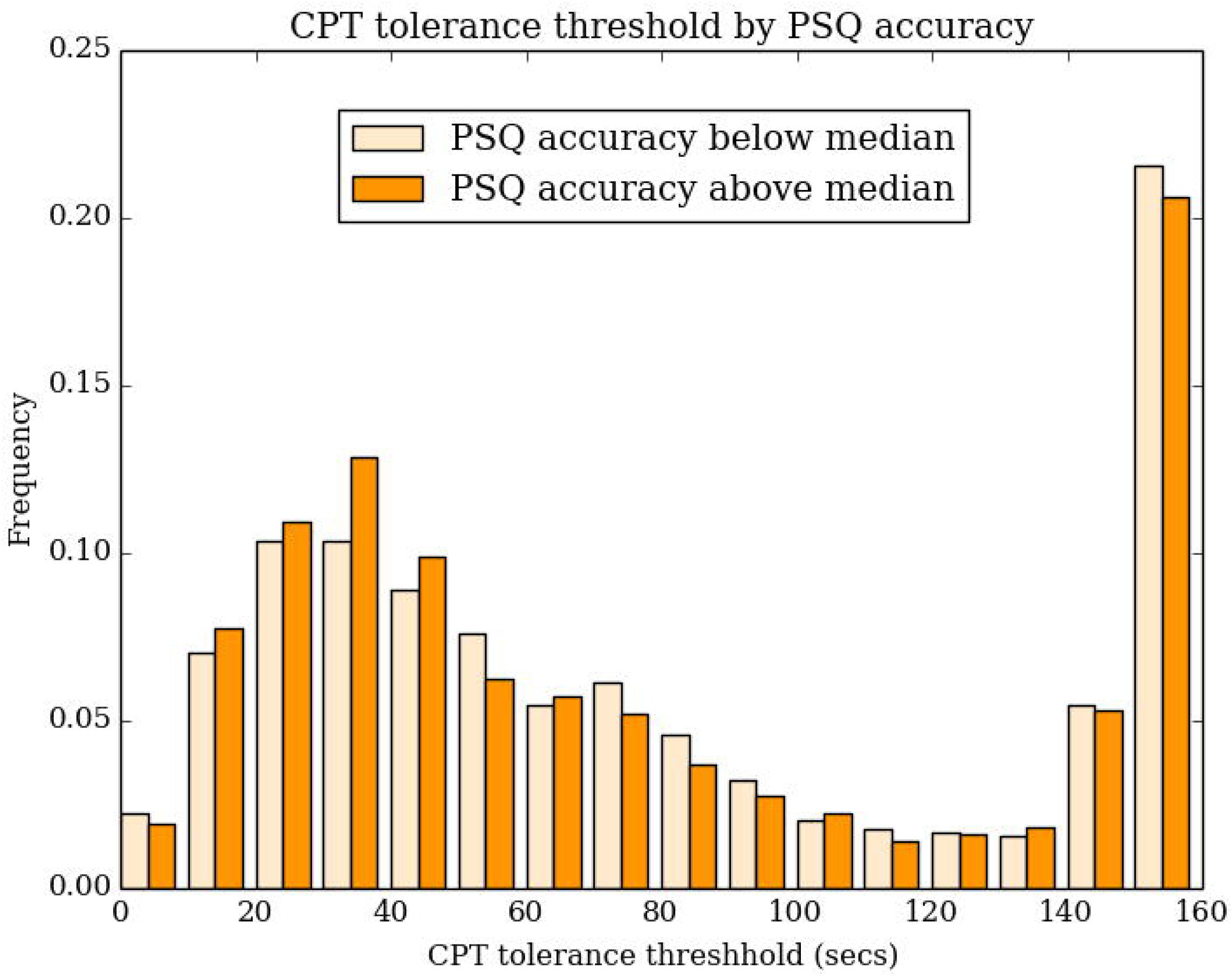
Distributions of CPT tolerance times by PSQ rating accuracy above or below the median among 2,107 participants who completed both PSQ and CPT.

PSQ total score was somewhat less correlated with retrospective pain intensity rating just after the CPT (r = 0.30 [95% CI: 0.26, 0.34], Spearman rho = 0.30) than found by Ruscheweyh et al. ^16^ who asked about pain intensity during the task (r = 0.56), but more similar to the correlation found in the validation of the Norwegian-language PSQ (r = 0.36). ^25^ Conversely, Ruscheweyh et al. ^16^ found no significant correlation of PSQ total with CPT pain threshold (r = 0.03, p = 0.86, n = 47), whereas we found small but significant correlations with both time to the first report of pain in the CPT (r = -0.14 [-0.19, -0.09], Spearman rho = -0.15) and time to withdrawal of the hand, or “tolerance”, (r = -0.22 [-0.27, -0.17], Spearman rho = -0.22). Ruscheweyh et al. ^16^ found no significant correlation of PSQ total with CPT pain threshold (r = 0.03, p = 0.86, n = 47), whereas the validation of the Norwegian-language PSQ ^25^ found a somewhat stronger correlation (r = -0.30, p<0.05, n=48).

469 participants (25%) reported maximum, retrospective pain intensities during the CPT below 3 on a scale of 0-10, where 0 is no pain at all and 10 is the worst pain imaginable. While we hypothesized that these participants might have prepared the CPT test incorrectly, they also reported lower PSQ total score (mean 2.55, SD 1.19 among those with CPT pain rating <= 3.0, mean 3.29, SD 1.35 among those with pain rating > 3.0, t=11.3, df=900.5, p<0.0001). Removing these participants from the analysis did not substantially change associations between the CPT and PSQ total score. The largest change after removal of these participants was a decrease in the correlation between CPT pain threshold and PSQ total score (from r = -0.14 [-0.19, -0.09] to r = -0.10 [-0.15, -0.05]).

### Accuracy

The 14 painful items of the PSQ can be ordered and weighted according to their painfulness. We therefore analyzed how accurately participants were to assign pain scores to the individual items. Within-person correlations between PSQ item ratings and loadings on the first principal component had a median of 0.62 (IQR: 0.31 - 0.81), with a long-left tail stretching into negative correlations. Accuracy scores showed small but significant correlations with both PSQ-total score (Spearman r = 0.14, p<0.0001) and CPT tolerance (Spearman r = -0.08, p=0.0002), suggesting that pain sensitivity and accuracy of pain rating related.

## Discussion

We measured pain sensitivity in a large population and an uncontrolled setting, where subjects answered the PSQ and performed a CPT at home. A limitation of the study is that we did not directly compare the results of self-administration of the CPT to results from a laboratory test within the same cohort. Instead, we compared the results of self-administration, in aggregate, to results reported in laboratory tests. For the critical measures of pain sensitivity, including cold pressor test tolerance, our results are similar to results obtained in controlled laboratory settings. Because of self-administration at home we had to adapt precise CPT protocol descriptions like temperature to simpler terms like room temperature. This also includes the amount of ice, which we described by 1/8^th^ of the water volume. Overall, this likely lead to considerable variability in the actual stimulus that was received by each participant but allowed us to conduct a web-based instruction workflow to self-administer a CPT in the absence of trained staff.

The online PSQ performed similarly to the original German-language version and sample, and to other versions and sample. The average PSQ-total score that we observed (3.3) was somewhat lower than that measured in small studies of several translation and in samples with and without chronic pain conditions ^3,9,17,18^, but a population study of 4,979 German-speaking Italians ^10^ also observed and average pain score of 3.3.

While we find strong psychometric support for the PSQ-total score, we show only weak replication of distinct minor and moderate PSQ scales, relative to a single PSQ scale. In this study, we see stronger overall evidence for a one-factor than a two-factor solution of the PSQ, consistent with results from the large study of the German-speaking Italians ^10^. The PSQ factor structure that we observed was similar to that seen for the Polish and French-language versions of the PSQ, which showed weak distinction between the minor and moderate pain factors, particularly for questions 3, 6, 7, 8, 16, and 17 ^3,9^. The Polish-language version was validated in a sample of 161 lower back pain patients, and the French-language version in two samples, one of 146 pre-surgical patients and the other of 85 health controls. The factor structure for the English-language PSQ has not been reported previously ^18^.

Among the participants who completed both the PSQ and an online self-administration of the CPT, the distribution of CPT scores was largely within the rather broad range of those found in laboratory studies, with the exception of more frequent intermodal thresholds in the online cohort. Whereas Ruscheweyh et al. ^16^ found no significant association between PSQ and CPT thresholds, we found small but significant negative correlations between PSQ score and both time to report of pain and to removal of the hand. The original validation study may have been underpowered to detect these associations. Power to detect the correlation of 0.22 in a sample of 47 is only 32%. Alternatively, participants who report lower pain sensitivity might over-report their pain threshold in the online design, but not in a laboratory design. Future studies should directly compare CPT performance in laboratory and self-administered settings.

These results support the notion that, when properly instructed, subjects are capable of self-administering the CPT in the absence of trained staff providing individual instructions. While variation in the how participants prepare the test apparatus, for example, the temperature of the water ^11^, certainly affect the reported pain thresholds, our results suggest that these errors will not yield false-positive associations with factors of interest to researchers. In fact, earlier studies have already shown that web-based phenotyping can produce a phenotype similar enough to physician-obtained phenotypes to yield similar results in genome-wide association studies ^24^. Our study adds to this in showing that web-based, self-phenotyping appears to be a valid approach not only when considering questionnaires, but that some test procedures can be followed to yield results similar to results from laboratory settings. Since we are using a web-based approach we do not have a familiarization session at the beginning which is usually done in a supervised laboratory test like QST. Instead we implemented a dry test before the actual time recordings started so that the participants were prepared to use the web interface and knew about the upcoming sequential steps.

In addition to supporting the validity of subjective and self-administered measures of pain sensitivity, our results also suggest that individuals differ in their ability to precisely and accurately rate pain, the latter measured by consistency with the observed factor structure, which aligns with other recent research ^20,21^. However, apparent differences might also be explained by, for example, differences in general attentiveness or conscientiousness, rather than pain rating ability as such. Accuracy scores had a long-left tail stretching into negative scores (i.e. scoring less painful items as more painful). Such a left tail is also observed in measures of person fit to item response theory models of the participant’s knowledge of the correct answers to a set of questions. These have been interpreted to reflect, for example, lack of attention to the task, rather than lack of ability, which is thought to have a more symmetric distribution ^4^. Factors like attentiveness are difficult to assess in a non-supervised setting like the one used for this study.

The online implementation, and related study design compromises, have several disadvantages. As we recruited participants via an online platform throughout the United States, it was impractical for us to repeat the same study with the same cohort under laboratory conditions. Therefore, we could not directly compare our results within the same cohort under controlled conditions but instead used data from published cohorts. We anticipate future studies directly comparing online and lab implementations.

Population-based, multi-omics studies will grow in the future and therefore a deep phenotypic characterization of datasets is key to deriving maximum amount of insight out of these efforts. Our approach of at-home testing and questionnaires is a first attempt to fill this gap. An online approach, by broadening participation in multi-omics studies, might help to further fill the gap of understanding pain phenotype subgroups, and discovering new pain-related biological pathways.

Upcoming investigations will include the identification of patients whose pain perception is different from the average, especially those able to discriminate better between more and less painful stimuli. Furthermore, we will continue to investigate additional pain testing@home possibilities and will identify pain relevant genes and pathways in order to derive targets using genetic association studies to relevant pain measures. This may allow us to adjust pain scales to standardize individual responses and to identify groups of patients for precision medicine approaches.

## Data Availability

All data is contained in the manuscript and supplementary material

## Acknowledgments

This research was conducted by 23andMe and Grünenthal GmbH. All authors are employees of these companies. We thank the research participants of 23andMe for making this study possible.

## Supplementary data

High resolution pictures of the web-based version of the CPT in addition to Figure 1 are available (supplementary Figures)

